# Deep Learning Unlocks the True Potential of Organ Donation after Circulatory Death with Accurate Prediction of Time-to-Death

**DOI:** 10.1101/2024.11.07.24316924

**Authors:** Xingzhi Sun, Edward De Brouwer, Chen Liu, Smita Krishnaswamy, Ramesh Batra

## Abstract

Increasing the number of organ donations after circulatory death (DCD) has been identified as one of the most important ways of addressing the ongoing organ shortage. While recent technological advances in organ transplantation have increased their success rate, a substantial challenge in increasing the number of DCD donations resides in the uncertainty regarding the timing of cardiac death after terminal extubation, impacting the risk of prolonged ischemic organ injury, and negatively affecting post-transplant outcomes. In this study, we trained and externally validated an ODE-RNN model, which combines recurrent neural network with neural ordinary equations and excels in processing irregularly-sampled time series data. The model is designed to predict time-to-death following terminal extubation in the intensive care unit (ICU) using the last 24 hours of clinical observations. Our model was trained on a cohort of 3,238 patients from Yale New Haven Hospital, and validated on an external cohort of 1,908 patients from six hospitals across Connecticut. The model achieved accuracies of 95.3 ± 1.0% and 95.4 ± 0.7% for predicting whether death would occur in the first 30 and 60 minutes, respectively, with a calibration error of 0.024 ± 0.009. Heart rate, respiratory rate, mean arterial blood pressure (MAP), oxygen saturation (SpO2), and Glasgow Coma Scale (GCS) scores were identified as the most important predictors. Surpassing existing clinical scores, our model sets the stage for reduced organ acquisition costs and improved post-transplant outcomes.

## Introduction

Organ donation plays a critical role in saving lives and improving the quality of life for individuals suffering from end-organ failure. Historically, organs from donation after brain death (DBD) donors have constituted the predominant source of transplantable organs, with donation after circulatory death (DCD) contributing to a comparatively smaller, albeit recently increasing, fraction^1^. A major reason for this disparity is the lower organ-yield from DCD donors, due to the reduced quality and longevity of allografts^2^. However, in the last 5 years, technological explosion of normothermic machine perfusion (NMP) and normothermic regional perfusion (NRP) have improved organ quality from DCD donors, highlighting the unrecognized potential of DCD donors in the transplant community^3,4^. Given these recent advances, there is now a growing consensus that augmenting DCD practice represents the largest and underutilized opportunity for expanding the organ donor pool^5^.

Although NMP and NRP work to improve the quality of organs procured from a DCD, the critical challenge limiting the volume of DCD practice is the unpredictability regarding whether, or when, a patient after terminal extubation (TE) will progress to meet the Uniform Declaration of Death Act (UDDA)^6^ criteria for organ donation. This uncertainty limits the ability of Organ Procurement Organizations (OPOs) to evaluate a potential DCD donor for organ donation and thus negatively impacts the organ yield from DCD donors. Indeed, while conventional guidelines stipulate that circulatory death must occur within a narrow time-frame following the cessation of life-sustaining treatment, only 59-72% of potential DCD donors die within the first hour^1,7^. The goal of this study is to investigate the potential of advanced machine learning models to accurately predict time-to-death (TTD) after extubation.

Recognizing the complexities inherent in DCD, we trained and externally validated an ODE-RNN model, which combines recurrent neural network with neural ordinary equations and excels in processing irregularly-sampled time series data. The model is designed to predict the time-to-death (TTD) of a patient following terminal extubation in the intensive care unit (ICU), leveraging the last 24 hours of clinical observations. Our model shows remarkable accuracy and calibration, underscoring its ability to accurately and reliably identify viable DCD organ donors, and enables a nuanced balance between the risks and benefits of a specific organ donation procedure.

A key challenge of modeling ICU data for TTD prediction is that the data consist of both static variables and a multidimensional time series of longitudinal variables, are measured at irregularly-spaced time points, and contain missing measurements in many variables. Previous efforts in predicting circulatory death within specified time frames include clinical risk scores such as the United Organ Sharing (UNOS) criteria^8^, and machine learning models such as XGBoost^9^, RNN^10^, LSTM^11^, GRU^12^, GRU-D^13^. However, these studies are predominantly based on conventional statistical models or basic machine learning architectures designed for regularly-sampled fixed-dimensional data. As a result, they cannot take full advantage of the data available, leading to insufficient performance and lower clinical reliability^7,14–17^. The UNOS criteria and XGBoost, a tree-based machine learning model, only consider static variables and cannot use the rich history of longitudinal variables. Recurrent neural network models and their extensions (RNN, LSTM, GRU and GRU-D) are able to model the time series of longitudinal variables, but they are primarily designed for data regularly-measured in time and perform badly on irregularly-sampled data. In contrast, we recommend using ODE-RNN, an architecture that builds upon recent advances in longitudinal modeling through the use of neural ordinary differential equations^18–20^, which specifically address the challenges posed by the irregularly-sampled data. The proposed model also allows one to form *patient phenoscape* visualizations for a better understanding of the cohort’s structure and heterogeneity. By leveraging state-of-the-art deep learning and representation learning methodologies, our approach surpasses the limitations of previous models and sets the stage for a more accurate and clinically relevant prediction of time-to-death following extubation, thereby promising an increase in the DCD donor pool.

## Results

### Modeling Longitudinal Clinical Variables Acquired at Irregular Time Intervals

Our model uses the last 24 hours of clinical observations of a patient prior to terminal extubation, which contains both static and longitudinal variables. The latter pose challenges for statistical analysis and machine learning methods due to their irregular measurements over time and the presence of missing values. We therefore used an Ordinary Differential Equation Recurrent Neural Network (ODE-RNN)^18^, a recent state-of-the-art deep learning architecture that addresses both issues. ODE-RNN integrates a recurrent neural network (RNN)^10^ component tailored for sequential modeling, with a Neural ODE^21^ component that interpolates between irregularly-sampled time points.

The model operates by accumulating longitudinal variables with static variables to create a summary of the clinical history of each patient, that we call the latent phenotype^22^. The latent phenotype is then used by a classifier to predict patient outcomes (i.e. TTD). This procedure makes the ODE-RNN particularly effective at processing EHRs containing both static and longitudinal variables^23^. A graphical depiction of the architecture is presented in Figure 1. Further details of the model are described in the Methods section.

**Figure 1.**
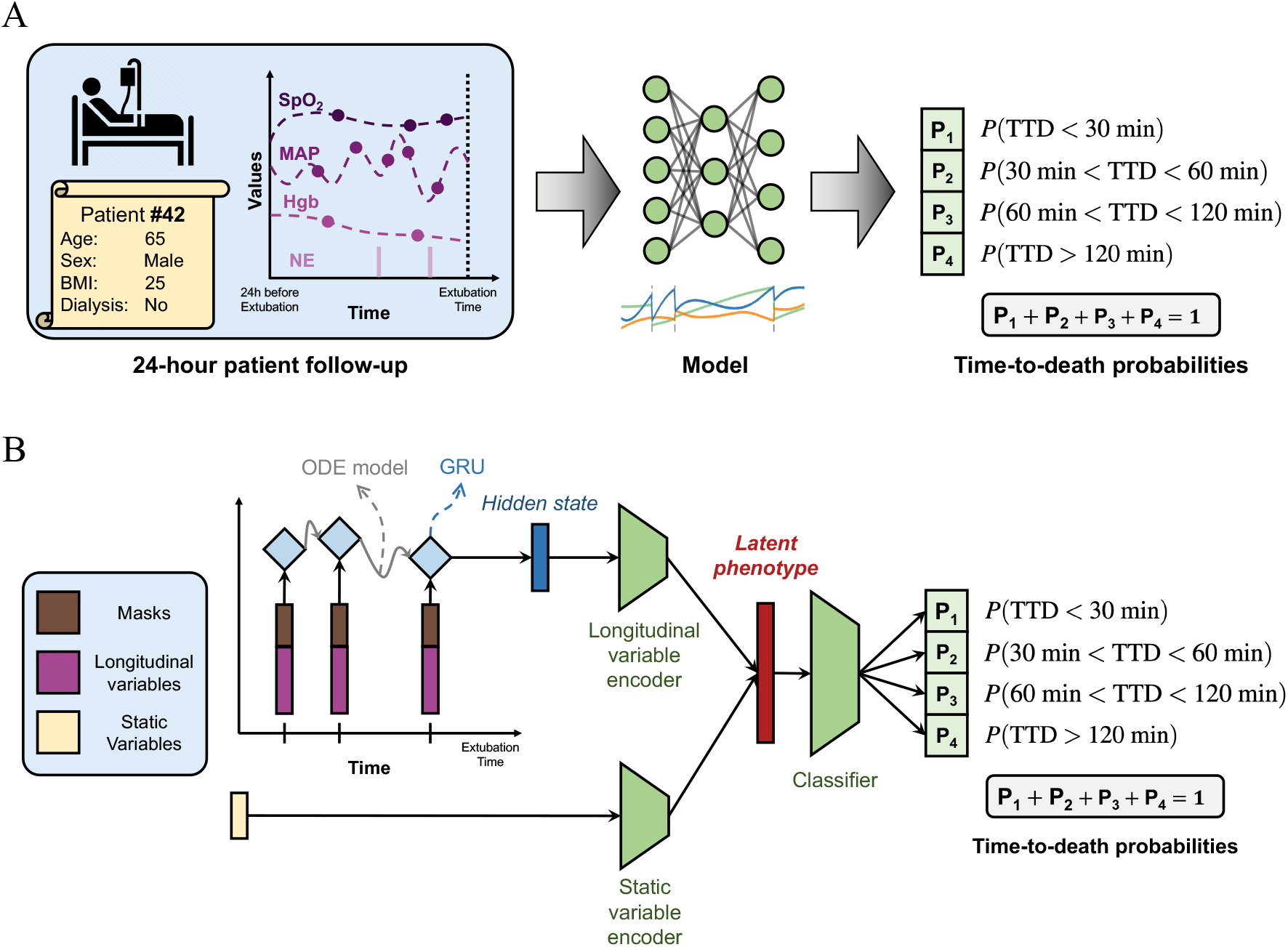
Description of the problem setup and model architecture. **A**. Problem setup. Based on the static variables of a specific patient (e.g. age, sex) and the last 24 hours of clinical follow-up prior to extubation (e.g. SpO2, MAP), our model predicts 4 probabilities: the probability that the time-to-death (TTD) is shorter than 30 minutes, between 30 and 60 minutes, between 60 and 120 minutes, and longer than 120 minutes. The sum of these probabilities equals to 1 by design. BMI stands for body mass index, SpO2 for oxygen saturation, MAP for mean arterial blood pressure, Hgb for hemoglobin, and NE for norepinephrine. Note that we consider 5 static variables and 25 longitudinal variables, and only some are shown for illustration purposes. **B**. Architecture of our ODE-RNN. The set of variables fed to the model consists of a concatenation of the longitudinal variables available at that observation time and a mask specifying which longitudinal variables are observed. Each clinical observation is sequentially processed by a gated recurrent unit (GRU) that incorporates the observation into the hidden state representation from the previous samples in the time series. Between observations, an ordinary differential equation (ODE) models the evolution of the patient’s hidden state continuously over time, which enables processing of variable temporal intervals between subsequent observations. The hidden state obtained after the whole time series is then complemented with the static variables to form the latent phenotype, a vector representation that summarizes the whole available information about the patient. The end classification is performed by using a multi-layer perceptron classifier (MLP) that predicts the TTD probabilities from the latent phenotype.

### Predictive Performance Evaluation

We compared our method with the UNOS criteria, the most widely used clinical score for identifying DCD candidates^8^, and existing machine learning models that have been used for the prediction of clinical outcomes, including RNN^10^, LSTM^11^, GRU^12^, GRU-D^12^, and XGBoost^9^.

All machine learning models were trained on the Yale New Haven Hospital (YNHH) cohort using a temporal data split. Data from patients before 2021 was used for training the models. Patients after 2021 were used for evaluation only to ensure robustness to distribution shifts over time.

The models were trained to predict time-to-death as a categorical variable, i.e., whether TTD fell within a given time frame (0-30 min, 30-60 min, 60-120 min, or >120 min). We evaluated the different models according to the overall categorical accuracy as well as pairwise binary classification for different grouped time frames (e.g., <30 min vs. >30 min). For these binary groupings, we also computed the area under the positive and negative predicted values (PPV, NPV), the area under the receiver operating characteristic curve (AUC-ROC) and the area under the precision-recall curve (AUC-PR). To assess the calibration of the models, that is, how well the predicted values represent the true likelihood, we computed the expected calibration error (ECE)^24^.

Table 1 displays the comparative performance of various models on the YNHH patient cohort after 2021 and the external validation cohort. We found that ODE-RNN based model consistently outperformed other methods on all metrics, for TTD prediction at 30, 60 and 120 minutes. We note the high performance of the model despite the stringent experimental setup (temporal split and external validation), highlighting the robustness of the method. ODE-RNN also shows the best calibration, suggesting the probability outputs of the model are very reliable.

**Table 1.** Comparison of the performance results of various machine learning models and statistical models on the Yale New Haven Hospital (YNHH) test cohort and external validation cohort. ROC-AUC stands for area under the receiver operating characteristic curve, PR-AUC stands for area under the precision-recall curve, ECE stands for expected calibration error. XGBoost and UNOS have zero standard deviation because there is no stochasticity in the training procedure.

| model | UNOS | XGBoost | RNN | LSTM | GRU | GRU-D | ODE-RNN |
| --- | --- | --- | --- | --- | --- | --- | --- |
| Yale New Haven Hospital Test Cohort (Temporal Split, After 2021) |  |  |  |  |  |  |  |
| Accuracy | 0.627 ± 0.000 | 0.831 ± 0.000 | 0.816 ± 0.014 | 0.861 ± 0.010 | 0.862 ± 0.021 | 0.856 ± 0.014 | **0.878 ± 0.007** |
| Accuracy (<30 vs. >30) | 0.627 ± 0.000 | 0.900 ± 0.000 | 0.922 ± 0.020 | 0.947 ± 0.009 | 0.948 ± 0.006 | 0.941 ± 0.005 | **0.955 ± 0.010** |
| Accuracy (<60 vs. >60) | 0.711 ± 0.000 | 0.928 ± 0.000 | 0.892 ± 0.012 | 0.939 ± 0.007 | 0.946 ± 0.010 | 0.941 ± 0.006 | **0.953 ± 0.003** |
| Accuracy (<120 vs. >120) | 0.779 ± 0.000 | 0.934 ± 0.000 | 0.903 ± 0.004 | 0.924 ± 0.006 | 0.930 ± 0.011 | 0.929 ± 0.012 | **0.942 ± 0.003** |
| ROC-AUC (<30 vs. >30) | 0.584 ± 0.000 | 0.962 ± 0.000 | 0.952 ± 0.023 | 0.978 ± 0.012 | 0.973 ± 0.010 | 0.972 ± 0.008 | **0.987 ± 0.004** |
| ROC-AUC (<60 vs. >60) | 0.592 ± 0.000 | 0.966 ± 0.000 | 0.932 ± 0.019 | 0.968 ± 0.012 | 0.965 ± 0.011 | 0.963 ± 0.007 | **0.987 ± 0.003** |
| ROC-AUC (<120 vs. >120) | 0.623 ± 0.000 | 0.975 ± 0.000 | 0.922 ± 0.018 | 0.961 ± 0.014 | 0.957 ± 0.011 | 0.951 ± 0.006 | **0.984 ± 0.002** |
| PR-AUC (<30 vs. >30) | 0.734 ± 0.000 | 0.972 ± 0.000 | 0.953 ± 0.034 | 0.981 ± 0.014 | 0.971 ± 0.012 | 0.975 ± 0.012 | **0.987 ± 0.003** |
| PR-AUC (<60 vs. >60) | 0.799 ± 0.000 | 0.986 ± 0.000 | 0.956 ± 0.024 | 0.982 ± 0.010 | 0.974 ± 0.010 | 0.976 ± 0.008 | **0.995 ± 0.001** |
| PR-AUC (<120 vs. >120) | 0.857 ± 0.000 | 0.993 ± 0.000 | 0.962 ± 0.019 | 0.985 ± 0.008 | 0.977 ± 0.010 | 0.974 ± 0.009 | **0.996 ± 0.001** |
| F1 (<30 vs. >30) | 0.770 ± 0.000 | 0.929 ± 0.000 | 0.940 ± 0.013 | 0.958 ± 0.007 | 0.960 ± 0.003 | 0.954 ± 0.003 | **0.967 ± 0.007** |
| F1 (<60 vs. >60) | 0.831 ± 0.000 | 0.949 ± 0.000 | 0.926 ± 0.008 | 0.957 ± 0.004 | 0.963 ± 0.006 | 0.959 ± 0.003 | **0.968 ± 0.002** |
| F1 (<120 vs. >120) | 0.876 ± 0.000 | 0.957 ± 0.000 | 0.935 ± 0.006 | 0.953 ± 0.003 | 0.957 ± 0.006 | 0.953 ± 0.009 | **0.960 ± 0.003** |
| PPV (<30 vs. >30) | 0.627 ± 0.000 | 0.882 ± 0.000 | 0.930 ± 0.024 | 0.942 ± 0.010 | 0.945 ± 0.010 | 0.935 ± 0.010 | **0.963 ± 0.010** |
| PPV (<60 vs. >60) | 0.711 ± 0.000 | 0.944 ± 0.000 | 0.922 ± 0.010 | 0.945 ± 0.007 | 0.952 ± 0.013 | 0.945 ± 0.008 | **0.967 ± 0.003** |
| PPV (<120 vs. >120) | 0.779 ± 0.000 | 0.966 ± 0.000 | 0.925 ± 0.015 | 0.937 ± 0.015 | 0.935 ± 0.018 | 0.927 ± 0.019 | **0.976 ± 0.010** |
| NPV (<30 vs. >30) | 0.000 ± 0.000 | **0.960 ± 0.000** | 0.915 ± 0.030 | 0.955 ± 0.018 | 0.956 ± 0.008 | 0.954 ± 0.009 | 0.953 ± 0.013 |
| NPV (<60 vs. >60) | 0.000 ± 0.000 | 0.886 ± 0.000 | 0.826 ± 0.023 | 0.922 ± 0.025 | **0.935 ± 0.006** | 0.928 ± 0.011 | 0.922 ± 0.011 |
| NPV (<120 vs. >120) | 0.000 ± 0.000 | 0.829 ± 0.000 | 0.797 ± 0.039 | 0.885 ± 0.042 | **0.919 ± 0.048** | 0.914 ± 0.025 | 0.827 ± 0.018 |
| ECE | 0.054 ± 0.000 | 0.092 ± 0.000 | 0.051 ± 0.004 | 0.048 ± 0.007 | 0.055 ± 0.014 | 0.055 ± 0.008 | **0.033 ± 0.008** |
| External Validation Cohort |  |  |  |  |  |  |  |
| Accuracy | 0.641 ± 0.000 | 0.785 ± 0.000 | 0.791 ± 0.015 | 0.800 ± 0.015 | 0.821 ± 0.010 | 0.809 ± 0.012 | **0.866 ± 0.010** |
| Accuracy (<30 vs. >30) | 0.641 ± 0.000 | 0.884 ± 0.000 | 0.923 ± 0.017 | 0.930 ± 0.003 | 0.942 ± 0.001 | 0.934 ± 0.003 | **0.953 ± 0.012** |
| Accuracy (<60 vs. >60) | 0.713 ± 0.000 | 0.898 ± 0.000 | 0.892 ± 0.014 | 0.913 ± 0.009 | 0.932 ± 0.005 | 0.919 ± 0.008 | **0.954 ± 0.007** |
| Accuracy (<120 vs. >120) | 0.786 ± 0.000 | 0.883 ± 0.000 | 0.865 ± 0.004 | 0.877 ± 0.012 | 0.892 ± 0.008 | 0.884 ± 0.011 | **0.934 ± 0.005** |
| ROC-AUC (<30 vs. >30) | 0.508 ± 0.000 | 0.943 ± 0.000 | 0.946 ± 0.012 | 0.964 ± 0.003 | 0.966 ± 0.004 | 0.965 ± 0.003 | **0.989 ± 0.005** |
| ROC-AUC (<60 vs. >60) | 0.506 ± 0.000 | 0.952 ± 0.000 | 0.928 ± 0.007 | 0.950 ± 0.005 | 0.955 ± 0.004 | 0.952 ± 0.003 | **0.987 ± 0.003** |
| ROC-AUC (<120 vs. >120) | 0.534 ± 0.000 | 0.945 ± 0.000 | 0.896 ± 0.005 | 0.915 ± 0.006 | 0.926 ± 0.004 | 0.923 ± 0.004 | **0.971 ± 0.004** |
| PR-AUC (<30 vs. >30) | 0.695 ± 0.000 | 0.961 ± 0.000 | 0.950 ± 0.018 | 0.971 ± 0.005 | 0.956 ± 0.015 | 0.967 ± 0.008 | **0.994 ± 0.003** |
| PR-AUC (<60 vs. >60) | 0.752 ± 0.000 | 0.978 ± 0.000 | 0.951 ± 0.012 | 0.972 ± 0.004 | 0.959 ± 0.012 | 0.967 ± 0.006 | **0.995 ± 0.001** |
| PR-AUC (<120 vs. >120) | 0.824 ± 0.000 | 0.985 ± 0.000 | 0.946 ± 0.007 | 0.968 ± 0.005 | 0.960 ± 0.004 | 0.963 ± 0.004 | **0.993 ± 0.001** |
| F1 (<30 vs. >30) | 0.781 ± 0.000 | 0.918 ± 0.000 | 0.937 ± 0.013 | 0.946 ± 0.002 | 0.955 ± 0.001 | 0.949 ± 0.003 | **0.966 ± 0.010** |
| F1 (<60 vs. >60) | 0.833 ± 0.000 | 0.932 ± 0.000 | 0.926 ± 0.007 | 0.941 ± 0.005 | 0.954 ± 0.003 | 0.945 ± 0.004 | **0.967 ± 0.004** |
| F1 (<120 vs. >120) | 0.880 ± 0.000 | 0.926 ± 0.000 | 0.913 ± 0.005 | 0.924 ± 0.006 | 0.933 ± 0.004 | 0.929 ± 0.007 | **0.955 ± 0.003** |
| PPV (<30 vs. >30) | 0.641 ± 0.000 | 0.871 ± 0.000 | 0.947 ± 0.013 | 0.937 ± 0.009 | 0.949 ± 0.005 | 0.937 ± 0.007 | **0.961 ± 0.009** |
| PPV (<60 vs. >60) | 0.713 ± 0.000 | 0.903 ± 0.000 | 0.925 ± 0.017 | 0.925 ± 0.018 | 0.938 ± 0.007 | 0.922 ± 0.010 | **0.958 ± 0.010** |
| PPV (<120 vs. >120) | 0.786 ± 0.000 | 0.905 ± 0.000 | 0.892 ± 0.016 | 0.891 ± 0.022 | 0.902 ± 0.016 | 0.890 ± 0.016 | **0.966 ± 0.011** |
| NPV (<30 vs. >30) | 0.000 ± 0.000 | 0.932 ± 0.000 | 0.876 ± 0.032 | 0.917 ± 0.013 | 0.928 ± 0.010 | 0.928 ± 0.015 | **0.949 ± 0.025** |
| NPV (<60 vs. >60) | 0.000 ± 0.000 | 0.889 ± 0.000 | 0.820 ± 0.015 | 0.886 ± 0.019 | 0.919 ± 0.008 | 0.910 ± 0.015 | **0.939 ± 0.010** |
| NPV (<120 vs. >120) | 0.000 ± 0.000 | 0.769 ± 0.000 | 0.713 ± 0.027 | 0.794 ± 0.020 | 0.836 ± 0.032 | **0.839 ± 0.013** | 0.815 ± 0.015 |
| ECE | 0.032 ± 0.000 | 0.111 ± 0.000 | 0.035 ± 0.008 | 0.085 ± 0.020 | 0.074 ± 0.007 | 0.074 ± 0.007 | **0.024 ± 0.009** |

The poor performance of XGBoost and UNOS can be explained by (1) the inability of UNOS criteria and XGBoost to capture the temporality of the patient’s data, i.e. they only use the last observation at the time of extubation; (2) the limited number of clinical variables used in UNOS (14 variables) compared to our ODE-RNN model (5 static and 25 longitudinal variables).

In Figure 2 panel B, we report the calibration plot of the ODE-RNN model for the binary prediction (< 30 min vs. > 30 min) on the external validation cohort. Calibration plots for other binary tasks are available in the Supplementary Materials. The model tended to give a reliable but conservative estimate of the probability of death within 30 minutes. Importantly, the model appeared well calibrated for low and high predicted probabilities, highlighting its reliability.

**Figure 2.**
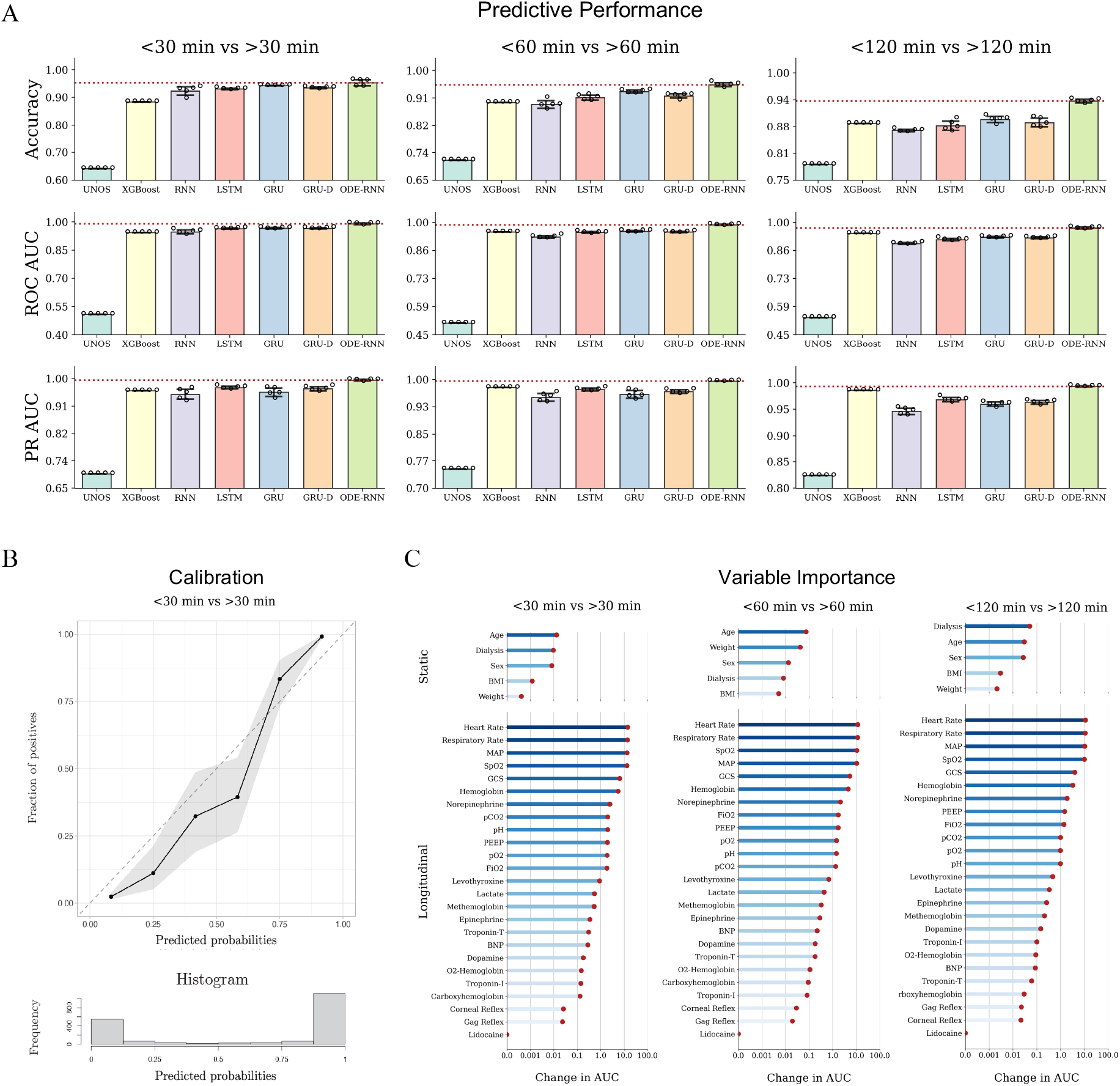
Model performances and analyses. **A**. Graphical representation of performance assessment for the UNOS score, XGBoost, LSTM, and ODE-RNN across different binary tasks. Left: TTD<30 min vs. TTD>30 min; Middle: TTD<60 min vs. TTD>60 min. Right: TTD<120 min vs TTD>120 min. We report the binary accuracy, the area under the receiver operating characteristic curve (ROC AUC), and the precision recall curve (PR AUC). ODE-RNN outperforms all other models for all tasks and all evaluation metrics. **B**. Calibration plot for the binary classification task TTD <30 min vs. TTD >30 min on the external validation cohort, computed with the R package val.prob.ci.2. The predicted probabilities come from the output of our model and plotted against the fraction of positives observed in the data. The histogram shows the prevalence of patients for different ranges of predicted probabilities. **C**. Variable importance for the predictions of the ODE-RNN model in the binary classification (left: <30 vs >30 minutes, middle: <60 vs >60 minutes, right: <120 vs >120 minutes). Variable importance was computed using permutation importance testing. The input variables were split into static variables (that do not change over time) and longitudinal variables. ROC-AUC stands for area under the receiver operating characteristic curves.

### Variable Importance Assessment

We assessed the importance and impact of the different clinical variables on the prediction of the models with permutation importance testing (Figure 2 panel C). For longitudinal variables, we found that heart rate was the most important variable in the prediction of the ODE-RNN, followed by respiratory rate, mean arterial blood pressure (MAP), oxygen saturation (SpO2), and the Glasgow Coma Scale (GCS) score. Corneal reflex and gag reflex which are frequently used by transplant surgeons and OPOs, appear to have the least impact on the prediction. Notably, static variables were found significantly less important than longitudinal variables, by an order of magnitude. We also found a strong consistency in the variable importance across different binary tasks.

### *Patient Phenoscape* Analysis

The patient phenotypes learned by our model enable accurate TTD predictions because they faithfully represent the patients’ condition and past clinical history. As such, these phenotypes provide richer information about the patients, compared to the single numerical value of TTD prediction. These phenotypes form a continuous landscape of the patient cohort, which we refer to as the *phenoscape*. Within the *phenoscape*, we can observe clusters of patients with similar conditions and continuous transitions from one condition to another, thereby uncovering the underlying dynamics of circulatory death. We used PHATE^25^, a dimensionality reduction method that preserves the underlyding data geometry^26–29^, to visualize the *phenoscape* and provide examples of new insights drawn from such analysis.

Our *patient phenoscape* visualizations in Figure 3 revealed that patients reside on a continuous spectrum of phenotype that goes beyond the TTD categorization. Patients were organized along an axis that corresponded with TTD but also with heart rate, SpO2, or GCS, among others. Finer investigation allowed us to identify the dynamical patterns most correlated with TTD, complementing the variable importance analysis above. For instance, in Figure 3 panel A, patients were colored according to their average heart rate and to their range of heart rate measurements (defined as the difference between highest and lowest values). While the range correlated with TTD, the average value did not, suggesting the variation in the heart rate is more important than the average.

**Figure 3.**
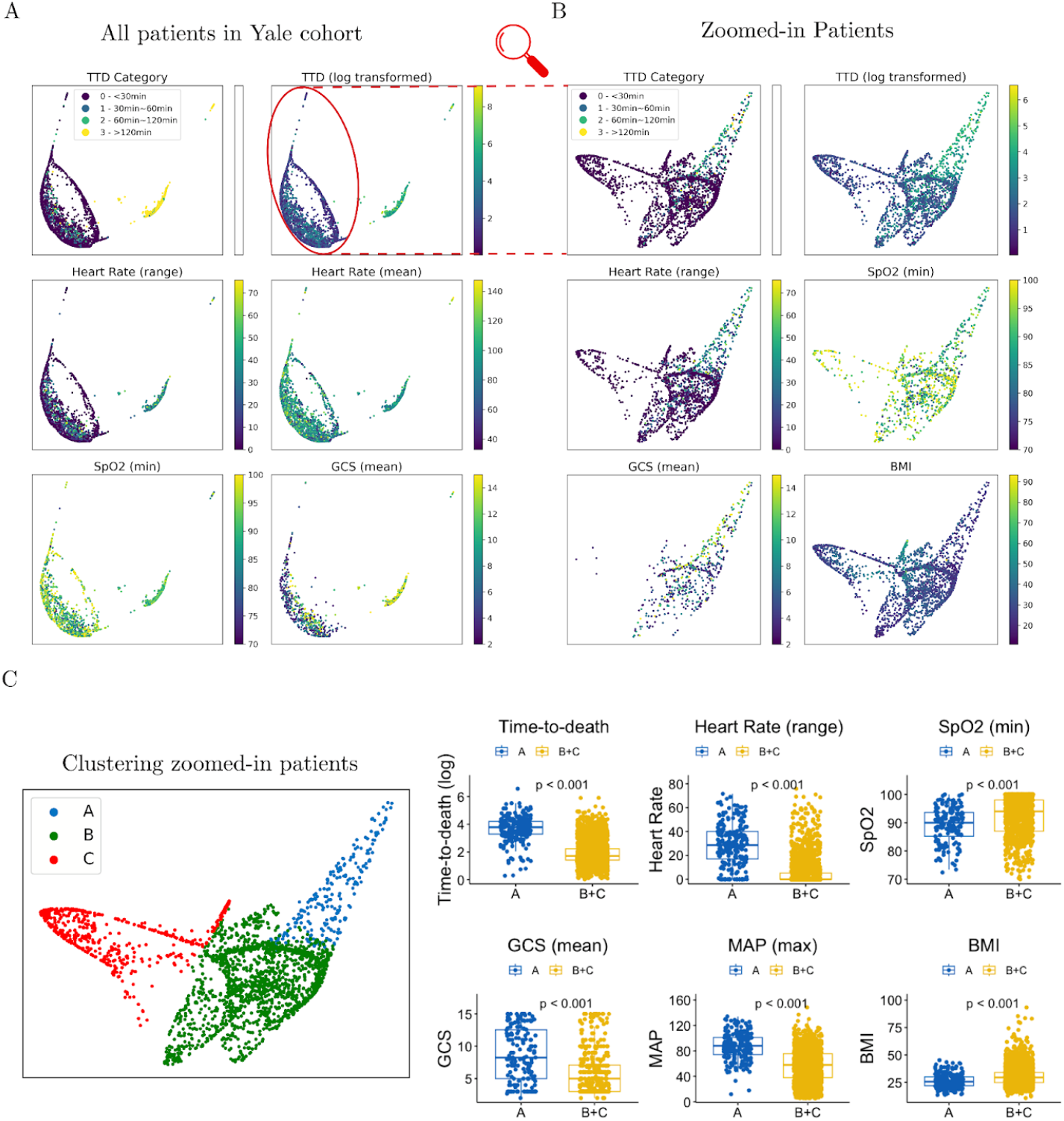
Visualization of the *patient phenoscape*. The latent phenotype is visualized in two dimensions using PHATE. In this plot each point represents a patient and the coloring is based on the value of different clinical variables. **A**. All patients in the Yale cohort are plotted and colored according to TTD label (top-left), TTD (log-transformed, top-right), heart rate (middle), SpO2 (bottom-left), and GCS (bottom-right). Each point represents a patient. These plots uncover the continuous structure of the patients’ latent phenotype and highlight the correlation between different clinical variables and the time-to-death. The longitudinal variables were transformed into scalar variables using different transformations. (range) computes the average of the five highest observations minus the average of the five lowest observations in the clinical history of the patient. (last) computes the average of the last five observations in the clinical history of the patient. (min) computes the average of the five lowest observations. Different transformations extract different patterns from the time series, enabling a finer interpretation of the dynamical patterns for a given clinical variable. For instance, we observed that heart rate (range) correlates with the TTD label, unlike heart rate (mean), suggesting the variation in heart rate is more important than the average value. The visualization of the whole cohort suggests two distincts groups of patients, characterized by high or low TTD. **B**. Focus visualization of the identified cluster of patients with TTD<120 min. This uncovers a finer grained structure in this specific cohort of patients. We colored the patients by TTD (log-transformed, top-left), range of heart rate (middle-left), minimum SpO2 (middle-right), average GCS (bottom-left), and BMI (bottom-right). **C**. We clustered the patients in the zoomed-in group of patients according to the similarity of their latent phenotype. We obtained three clusters: A, B, and C. Guided by the visualization of panel B, we examined the specific phenotype of patients from cluster A, which appear to have higher TTD than the rest of the patients. We show boxplots and corresponding independent t-tests p-values for difference of means between clusters, for various clinical variables. This analysis characterizes cluster A as a subgroup of patients with high TTD, high range of heart rate, low minimum SpO2, high GCS, as well as low BMI.

Our *phenoscape* visualization also showed a clear distinction between patients with TTD<120 minutes and another group with TTD>120 minutes (categories 0,1,2 vs. 3 in top-left of Figure 3 panel A). This separation suggests an obvious clinical difference between patients with TTD<120 min and TTD>120 min. We performed a fine-grained analysis of the former cluster, to identify clinical drivers that make the difference between short-range (≈30min) and medium-range (≈60min) TTD. Panels B and C in Figure 3 show a visualization of the identified group of patients with TTD<120min. From this zoomed-in analysis, we identified three clusters (A, B, and C). Guided by our visualization, we examined the specific phenotype of patients from cluster A, which appear to have higher TTD than rest of the patients. We found that patients from cluster A were characterized by higher TTD, higher range of heart rate, lower minimum SpO2, higher GCS, higher MAP and lower body mass index (BMI). This reveals that, over the last 24 hours before TE, range of heart rate, minimum SpO2, average GCS, and the maximum MAP observed are all predictive of TTD.

## Discussion

Augmenting the number of donations after circulatory death has been recognized as a crucial factor in mitigating the ongoing organ shortage, with the potential to increase the organ donor pool by as much as 30% in the United States^30^. However, a major factor hindering the rapid increase of DCD is the unpredictability regarding the time of circulatory death after extubation, leading to an unmanageable risk of prolonged warm ischemic injury. In the United Kingdom, it is estimated that 40% of donation teams mobilized for potential DCD donations are unsuccessful due to unpredictably long ischemic injury^31^. In the United States, only 59-72% of potential DCD donors die within the first hour after terminal extubation^1,7^. Similarly, in our cohort, only 73.8% of the patients died within the first hour.

This low success rate worsened by total unpredictability results in the waste of essential and valuable health-care resources and increased distress for families. Therefore, the average cost per DCD organ is estimated to be 63% higher than a DBD organ, mostly attributable to the unpredictability of DCDs from these “dry runs”^32,33^. The recent introduction of NMP and NRP, which have significantly improved the quality of organs from DCD by resuscitation of DCD organs prior to transplant; but that added expense has also contributed in making “dry runs” even more costly as the resources spent in mobilizing the NMP and NRP teams are still wasted on an unpredictable and thus failed DCD attempt^34,35^. Unsuccessful DCD donations also result in wasted human effort, and an avoidable environmental cost linked to the inherent logistics (air/ground transport) of a failed DCD attempt; and of an immeasurable psychological burden on grieving families hoping to make sense of their tragedy with the hope of a successful organ donation^36^.

These considerations highlight the importance and value of an accurate TTD prediction and have motivated the introduction of clinical scores, such as the UNOS criteria^8^ or the University of Wisconsin Donation after Circulatory Death evaluation tool (UW-DCD)^37^. However, these statistical methods show poor discrimination performance. For the UNOS criteria, PPV and NPV are reported to be 75.8% and 73%^38^. The UW-DCD, shows even worse performance (57.6% PPV and 61.8% NPV)^38^, and requires disconnecting the patient from ventilator for 10 minutes^37^. In contrast, our model achieved a PPV of 95.8%, a NPV of 93.9% and an accuracy of 95.4% for predicting whether the donor would die within the first hour on the external validation cohort, thereby only misclassifying 4.6% of the patients. Our model also does not require disconnecting the patient from ventilator, as seen in UW-DCD.

Predicting time to circulatory death has been previously attempted in the literature^8,14–17,37^. Nevertheless, previous studies predominantly have relied on conventional statistics and machine learning architectures such as logistic regression^7^ or long short term memory (LSTM)^11,14^. These models showed promising performance, but their simple architecture failed to fully capture the signal in data. Winter et al.^16^ proposed a model including only pediatric patients, with an AUC-ROC of 0.85, which is significantly lower than our model (AUC-ROC of 98.7 ± 0.3 for ODE-RNN). It is noteworthy that pediatric patients (<18 years) only conforms to 5% of total deceased organ donations in United States, whereas our model is applicable for the 95% organ donors in U.S.^39^. Furthermore, the performance evaluation in these studies was often limited and without calibration, preventing a thorough assessment of the maturity of models for a potential clinical use.

Our study aims at addressing these shortcomings by leveraging the most recent advances in machine learning and providing the most robust clinical evaluation possible. The ODE-RNN architecture is specifically designed to handle specific challenges of clinical time series, such as irregular sampling or missing data, which enables capturing all the relevant information in patient’s data. To evaluate the models as closely as possible to a realistic clinical practice scenario, we used temporal splitting, removing bias induced by a change of clinical practices over time. Notably, such an evaluation strategy was absent from previous works on TTD prediction. We also used an external validation cohort to remove the bias linked to the clinical practices at different hospitals.

In clinical setting, it is important that predictive models give a notion of certainty regarding their predictions. Indeed, having access to a probability of death within a timeframe is crucial in balancing the expected benefits and costs of a planned organ donation. Remarkably, we found that our model showed excellent calibration, suggesting that the predicted probabilities could be directly interpreted at face-value. Furthermore, we note that our model jointly predicts probabilities for all four time-frames (<30 min, 30∼60 min, 60∼120 min, >120 min), which enables a fine-grained evaluation. This facilitates work of OPOs and transplant centers, who plan procurement of particular organs based on their warm ischemia time acceptance criteria for that particular donor as standard or extended criteria.

Our variable importance analysis was generally consistent with the UNOS criteria variables with respiratory rate, heart rate, SpO2, PEEP, and norepinephrine ranking high. However, only dopamine, a variable in the UNOS model, was found to be one of the least important variables in our model. We also found that MAP and GCS, although absent from the UNOS criteria, were very important variables for the predictive accuracy in our model. It is noteworthy that, although UNOS model excludes both MAP and GCS, they have been previously identified as important predictors of death after TE^8,16^.

Deep learning architecture like, ODE-RNN, bring added value with ability to handle irregular time series of arbitrary length and to provide a hidden state representation of the patient: a latent phenotype. Our experimental results showed, the ability to process the whole available time-series, and handle irregular sampling, resulted in better predictive performance. We further showed that our model enables a fine-grained analysis of the patient cohort by producing a latent phenotype for each patient, put together visualized as a phenoscape. The phenoscape identifies and separates the specific subgroups of patients with higher TTD and could potentially support clinical discovery essential to the prediction and identification for a DCD donor.

While the model developed in this study represents an important proof-of-concept, showing compelling predictive performance, our study still suffers from several limitations. First, our patient cohorts were from various hospital records and not from the OPOs, therefore it is quite possible that some patients in our cohorts were ineligible for organ donation. Second, our model uses the last 24 hours before extubation of the patient and predicts TTD up to the time of extubation. This results from many patients in our cohort having a short follow-up time, preventing us from training a model that predicts TTD with more than 24 hours before extubation. We hope the exceptional performance of the model developed in this study will enable us to extend our work to a specific cohort of organ donors, which will enable us to have a longer follow-up and irrevocably demonstrate the utility of machine learning models for improving the success rate of DCD.

## Conclusions

The result of this study suggests that, dedicated state-of-the-art deep learning models can accurately and reliably predict time-to-death after terminal extubation, thereby overcoming a significant obstacle to increasing the number of successful DCDs. In addition, including the longitudinal clinical history of the patient was found to be crucial in achieving good performance. Future prospective studies will be needed to assess the exact gains in real-world clinical practice.

## Methods

### Patient cohort and data preparation

We used two separate cohorts of patients to develop and validate the model. The first cohort contained 3,238 patients at Yale New Haven hospital (YNHH) older than 18 years old, with a recorded TE in the ICU between 2014 and 2023. For unbiased validation, using same inclusion criteria, we formed an external cohort from six different hospitals with 1,908 patients. The hospitals from the external cohort are the Bridgeport hospital, Greenwich hospital, Lawrence and Memorial hospital, Saint Raphael hospital, Westerly hospital and the Yale New Haven Children’s hospital. The median time from extubation to death was 7.15 minutes in the first cohort and 8.28 minutes in the external cohort. The two cohorts are summarized in Table 2.

**Table 2.** Statistics of the two cohorts. BMI stands for body mass index, and TTD for time-to-death.

| Demographic Information | YNHH Cohort<br>(*n* = 3,238) | External Hospitals Cohort<br>(*n* = 1,908) |
| --- | --- | --- |
| Median Age, years | 68 | 74 |
| Sex, No. (%) |  |  |
| Male | 1,899 (58.1) | 1,081 (56.3) |
| Female | 1,372 (42.0) | 839 (43.7) |
| Median BMI | 28.44 | 28.50 |
| TTD, minutes |  |  |
| Median | 7.15 | 8.28 |
| Mean | 134.75 | 110.93 |
| Standard Deviation | 449.31 | 346.57 |
| Patients with TTD in range, No. (%) |  |  |
| 0~30 min | 2,156 (66.6) | 1,223 (64.1) |
| 30~60 min | 226 (7.0) | 138 (7.2) |
| 60~120 min | 217 (6.7) | 139 (7.3) |
| >120 min | 639 (19.7) | 408 (21.4) |

For each patient, we extracted 5 static variables and 25 longitudinal variables, collected up to 24 hours before extubation. Longitudinal variables were observed at a range of frequencies – from a single observation in 24 hours to one/minute. Missing values in the longitudinal records were imputed by performing a combination of forward-fill, backward-fill and mean-fill imputation^40^. In addition to imputation, presence of missing values was fed to the model by creating binary missingness indicators. TTD was defined as the time from terminal extubation to circulatory death. TTD was converted into an ordinal variable with 4 categories (0: 0∼30 minutes, 1: 30∼60 minutes, 2: 60∼120 minutes, and 3: longer than 120 minutes), that were used as target labels in the machine learning model. We chose these time frames as most transplant centers use 0-60 minutes as “standard criteria” for kidney DCD donation and 60*∼*120 minutes as an “extended spectrum”^41^; similarly, 0*∼*30 minutes as “standard criteria” for liver DCD donation and 30*∼*60 minutes as an “extended spectrum”^42^.

### List of clinical variables used in the models

The following longitudinal and clinical variables were extracted for each patient.

#### Longitudinal variables

B-type natriuretic peptide (BNP), carboxyhemoglobin, corneal reflex, fraction of inspired oxygen FiO2, gag reflex, Glasgow Coma Scale (GCS), hemoglobin, lactate, mean arterial blood pressure (MAP), methemoglobin, O2-Hemoglobin, partial pressure of carbon dioxide (pCO2), positive end-expiratory pressure (PEEP), blood potential of hydrogen (pH), partial pressure of oxygen (pO2), pulse, respirations, oxygen saturation (SpO2), Troponin-I, Troponin-T, Dopamine, Epinephrine, Levothyroxine, Lidocaine, Norepinephrine.

#### Static variables

Age, Body Mass Index (BMI), Dialysis, Sex, Weight.

### Model description

To capture the impact of longitudinal variables on the target while accommodating for the irregular sampling of clinical trajectories, we used an Ordinary Differential Equation Recurrent Neural Network (ODE-RNN)^18^. ODE-RNNs combine two powerful architectures, recurrent neural networks (RNN) and neural ordinary differential equations (Neural ODE), making them exceptionally apt at processing clinical time series^19,23^. RNNs are neural networks specialized for processing sequences. At each time step, they maintain a hidden state which represents the whole previous information in the time series. Upon reading a clinical record, the RNN updates its hidden state by combining the previous hidden state with the new observation, using an update unit (here a gated recurrent unit (GRU)). However, RNNs assume continuous time intervals between observations, an assumption typically not met in clinical time series. To address this limitation, ODE-RNN uses a Neural ODE, that describes dynamics in continuous time, to model the dynamics of the hidden state between observations. This uniquely allows the model to capture the full span of the clinical records of each patient, correctly accounting for the time interval between observations, and improving upon previous methods such as logistic regression, XGBoost, or the UNOS criteria, among others.

The model accumulates the last 24 hours before extubation of the patient’s vitals, medications used (e.g. vasopressors), neurological assessments, and lab results, together with their demographic records, and produces a representation of the patient, which we refer to as the patient’s latent phenotype. This latent phenotype can be understood as a learnt compact clinical summary of a particular patient. This representation is then used as an input to a multi-layer perceptron classifier that predicts the probability of each label category (0, 1, 2, or 3, corresponding to the 4 time ranges). Figure 1 depicts our model pipeline.

Our model contains the following neural networks:

- *f*_static_: a multi-layer perceptron (MLP) that processes the static variables.
- *f*_ODE_: an MLP that predicts the derivative of the hidden state dynamics between observations.
- *f*_GRU_: a gated recurrent unit (GRU) that updates the hidden states at each observation point.
- *f*_longitudinal_: an MLP that processes the final hidden state of the longitudinal variables.
- *f*_fusion_: an MLP that fuses the latent states derived from static and longitudinal variables, producing a latent variable called the *latent phenotype*.
- *f*_classifier_: an MLP that performs classification using the latent phenotype.

Algorithm 1 describes how the model predicts outcomes for a single patient. The model takes as input static variables *s* ∈ ℝ^*ℓ*^, longitudinal variables *x*_1_, …, *x*_*n*_ ∈ ℝ^*k*^, observation times *t*_1_, …, *t*_*n*_ ∈ ℝ, and boolean observation masks *m*_1_, …, *m*_*n*_ ∈ ℝ^*k*^, where *m*_*i*_ = (*m*_*i*1_, …, *m*_*ik*_). The value *m*_*ij*_ = true if *x*_*ij*_, the *j*^th^ variable at time *i*, is observed, and *m*_*ij*_ = false otherwise. Including these observation masks enables the model to utilize “informative missingness,” which is correlated with the patient’s condition. For instance, a patient is unlikely to be suspected of heart failure if B-type Natriuretic Peptide (BNP) is not frequently measured. The model iterates through the observation time points, updating the hidden state *h* of the longitudinal variables using *f*_ODE_ and *f*_GRU_. Between observation points, *f*_ODE_ is integrated to continuously update *h*, while at observation times, *f*_GRU_ updates *h* using *x*_*i*_, *m*_*i*_, *t*_*i*_. The final *h* contains accumulated information from the entire history of the longitudinal variables, which is then processed by *f*_longitudinal_ and fused with the processed static variables *s* (via *f*_static_) using *f*_fusion_. This results in a latent variable representing the patient’s condition, referred to as the *latent phenotype. f*_classifier_ uses the latent phenotype to make the final classification prediction.

#### Algorithm 1 ODE-RNN using GRU cell update (using one patient for illustration)

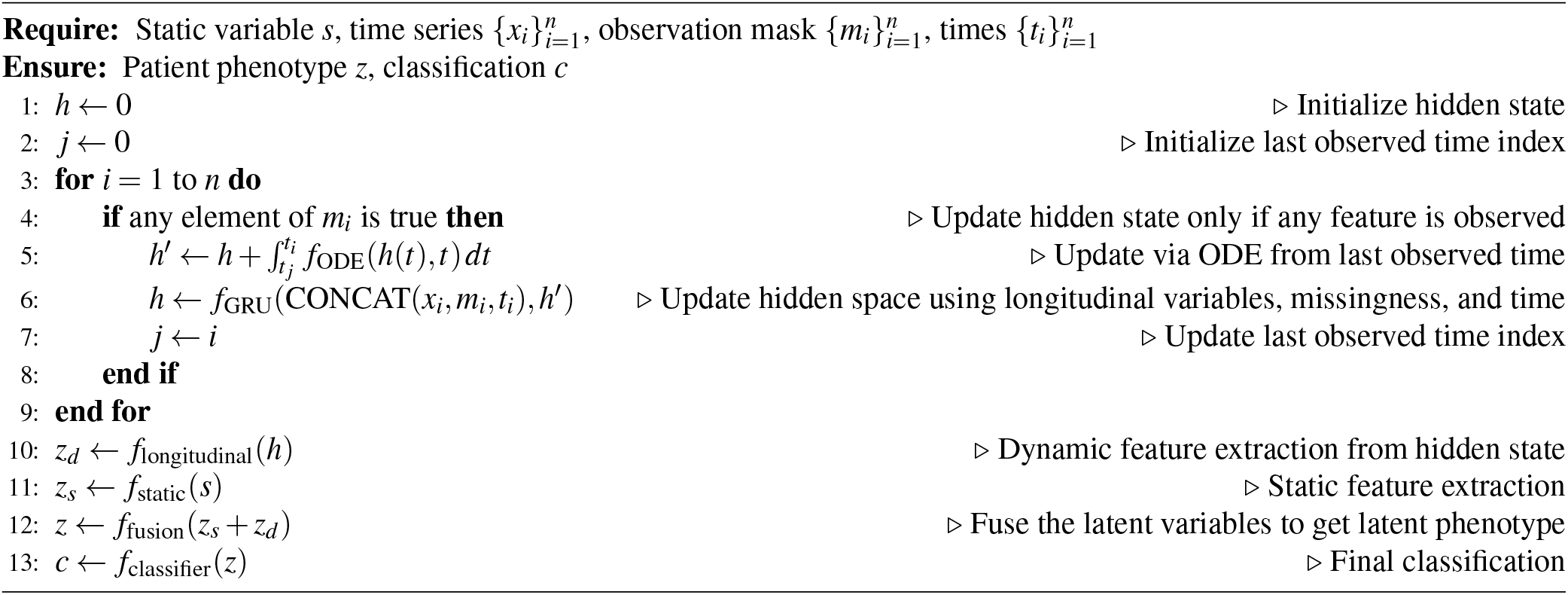

### Evaluation of model performance

We compared the performance of our approach with machine learning methods and clinical scores such as the UNOS criteria^8^. Machine learning methods directly learn associations from the available data while clinical scores consist of clinical criteria designed by experts. Within the machine learning methods, we distinguish between static methods and longitudinal methods. Unlike longitudinal methods, static methods cannot process a time series of clinical information. They are therefore trained on the last available clinical observation at the time of extubation only.

#### Static machine learning methods

1. **XGBoost** XGBoost^9^ is an effective and widely used tree-based machine learning algorithm for predictive modeling. Utilizing an ensemble of decision trees, XGBoost improves model accuracy by combining weak learners into a strong one. It is designed with sparsity awareness, which renders it helpful in clinical health data. However, being a static method, XGBoost cannot process longitudinal data.

#### Longitudinal machine learning methods

1. **RNN** Recurrent Neural Networks (RNNs)^10^ are a class of neural networks specialized for processing sequences, designed for handling time-series data or sequential information. A vanilla RNN processes sequences by iterating through elements, using its internal state to retain information from previous inputs.
2. **LSTM** Long Short-Term Memory (LSTM)^11^, an extension of vanilla RNNs, is designed to overcome the vanishing gradient problem by incorporating memory cells. These cells enable LSTMs to retain information over extended sequences, making them adept at tasks requiring longer-term dependencies.
3. **GRU** Gated Recurrent Units (GRUs)^12^ are a streamlined variant of LSTMs. Compared to the LSTM architecture, a GRU replaces the input, forget and output gates with the reset and update gates for higher efficiency.
4. **GRU-D** GRU-D, an extension of GRUs^13^, integrates decay mechanisms to handle missing data in time-series. It modifies the GRU architecture to accommodate irregularly-sampled data, enhancing prediction accuracy in such scenarios.

#### Clinical scores

Besides the machine learning models above, we also compared our approach to the most widely used clinical score for DCD candidates identification: the UNOS criteria^8^.

1. **UNOS** UNOS criteria consisted of fourteen clinical variables developed by the UNOS DCD consensus committee, based on expert opinion^8^. Criteria include physiological measurements (e.g. heart rate <30) and respiratory characteristics (e.g. FiO2 >0.5). A final score was computed by adding up the number of UNOS criteria present in the patient at the time of extubation.

For comparison, we trained these various models on the YNHH cohort using a temporal data split. Data from patients before 2021 was used for training the models. Patients after 2021 were used for evaluation only. This ensured that our results were robust to distribution shifts over time^43^. Standard errors were computed by training five different models with different initializations.

The models were trained to predict TTD as a categorical variable within a given time frame (0∼30 min, 30∼60 min, 60∼120 min, or >120 min). We evaluated the different models according to the overall categorical accuracy as well as pairwise binary classification for different grouped time-frames (e.g., <30 min vs. >30 min). For these binary groupings, we also computed the positive predictive values (PPV), negative predicted values (NPV), area under the receiver operating characteristic curve (AUC-ROC) and the area under the precision-recall curve (AUC-PR). The ROC curve measures the trade-off between sensitivity and specificity, while the PR curve measures the trade-off between precision and recall. To assess the calibration of the models, we computed the expected calibration error (ECE).

### Visualizing structures in high-dimensional patient phenoscape with PHATE

To predict TTD, our ODE-RNN model produces a latent phenotype for each patient, which can be intuitively understood as a learnt summary of the clinical history of the patient20. We explored the space of latent phenotypes of all patients in the cohort, the patient phenoscape, showing its potential to provide new clinical insights.

The latent phenotype of each patient is high-dimensional, and thus cannot be directly visualized. Therefore, we first produced a two-dimensional representation of the phenotype using PHATE^25^, a non-linear dimensionality reduction and visualization method that stays faithful to the geometry of the data and retains the inherent similarity between patients. In this visualization, each patient is represented as a point in a two-dimensional phenotypic space. The patient phenoscape is the set of the representations of all patients in the cohort. We colored each point according to their TTD and the value of certain clinical variables, enabling a fine-grained exploration of the impact of different clinical factors on the TTD.

## Data Availability

We are not going to make the data available to the general public, protecting the privacy of patients, and sensitive nature of the data in line with HIPPA rules.

## Acknowledgements

This research was funded and supported by NIH grants (1F30AI157270-01, R01HD100035, R01GM130847, R01GM135929), NSF Career grant 2047856, the Chan-Zuckerberg Initiative grants CZF2019-182702 and CZF2019-002440, 2020 Yale Innovation Grant, the Sloan Fellowship FG-2021-15883, and the Novo Nordisk grant GR112933. The content provided here is solely the responsibility of the authors and does not necessarily represent the official views of the funding agencies. The funders had no role in study design, data collection and analysis, decision to publish, or preparation of the manuscript.

## Author contributions statement

S.K. and R.B. identified the research problem. R.B. provided the data. X.S., E.D.B. and C.L. conceived the experiments. X.S. and E.D.B. conducted the experiments. X.S., E.D.B. and C.L. analyzed the results. S.K. and R.B. provided advice and supervision. All authors wrote and reviewed the manuscript.

## Supplementary Materials

### Calibration plots

**Figure 4.**
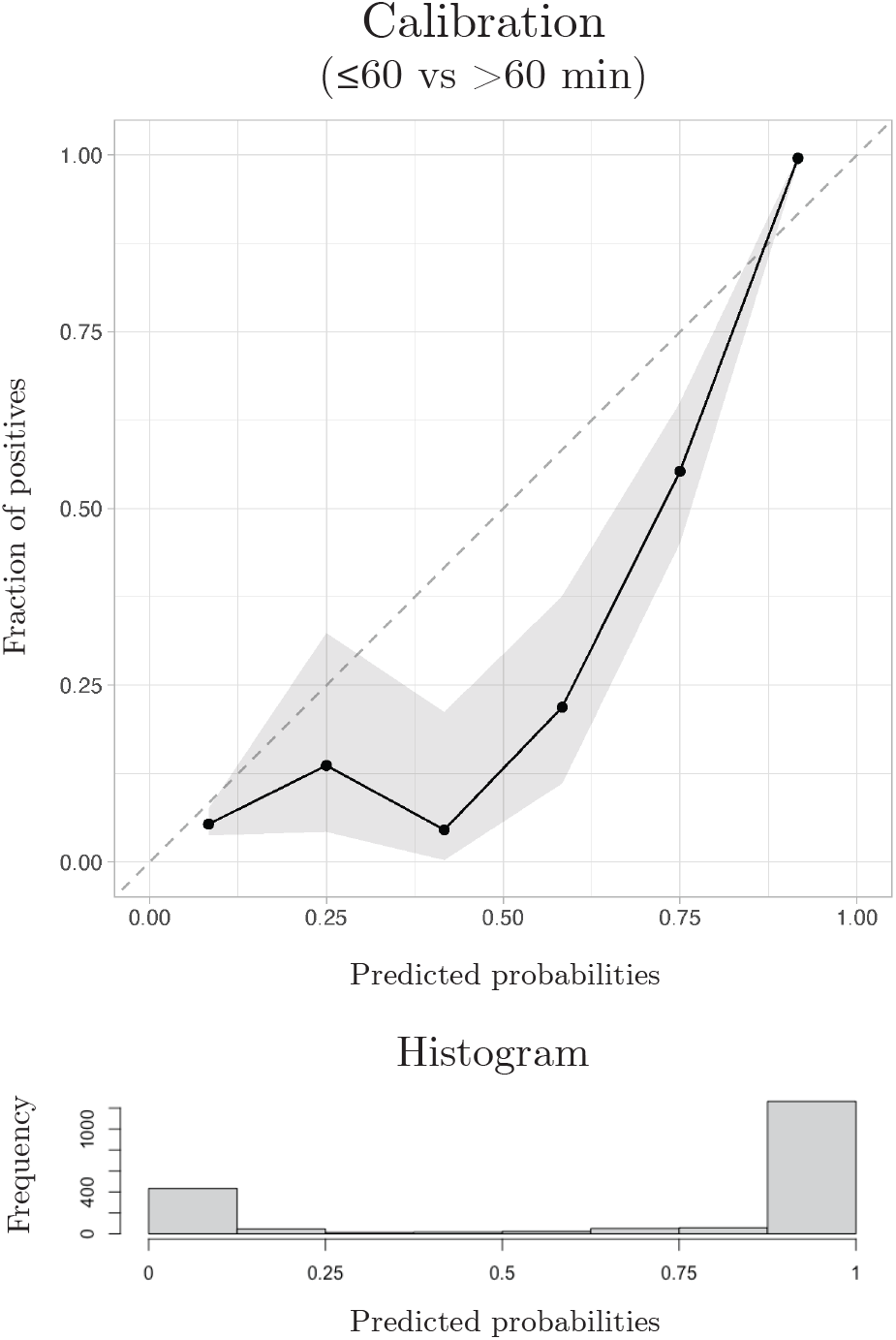
Calibration plot for the binary classification task TTD <60 min vs. TTD >60 min on the external validation cohort, computed with the R package val.prob.ci.2. The predicted probabilities come from the output of our model and plotted against the fraction of positives observed in the data. The histogram shows the prevalence of patients for different ranges of predicted probabilities.

**Figure 5.**
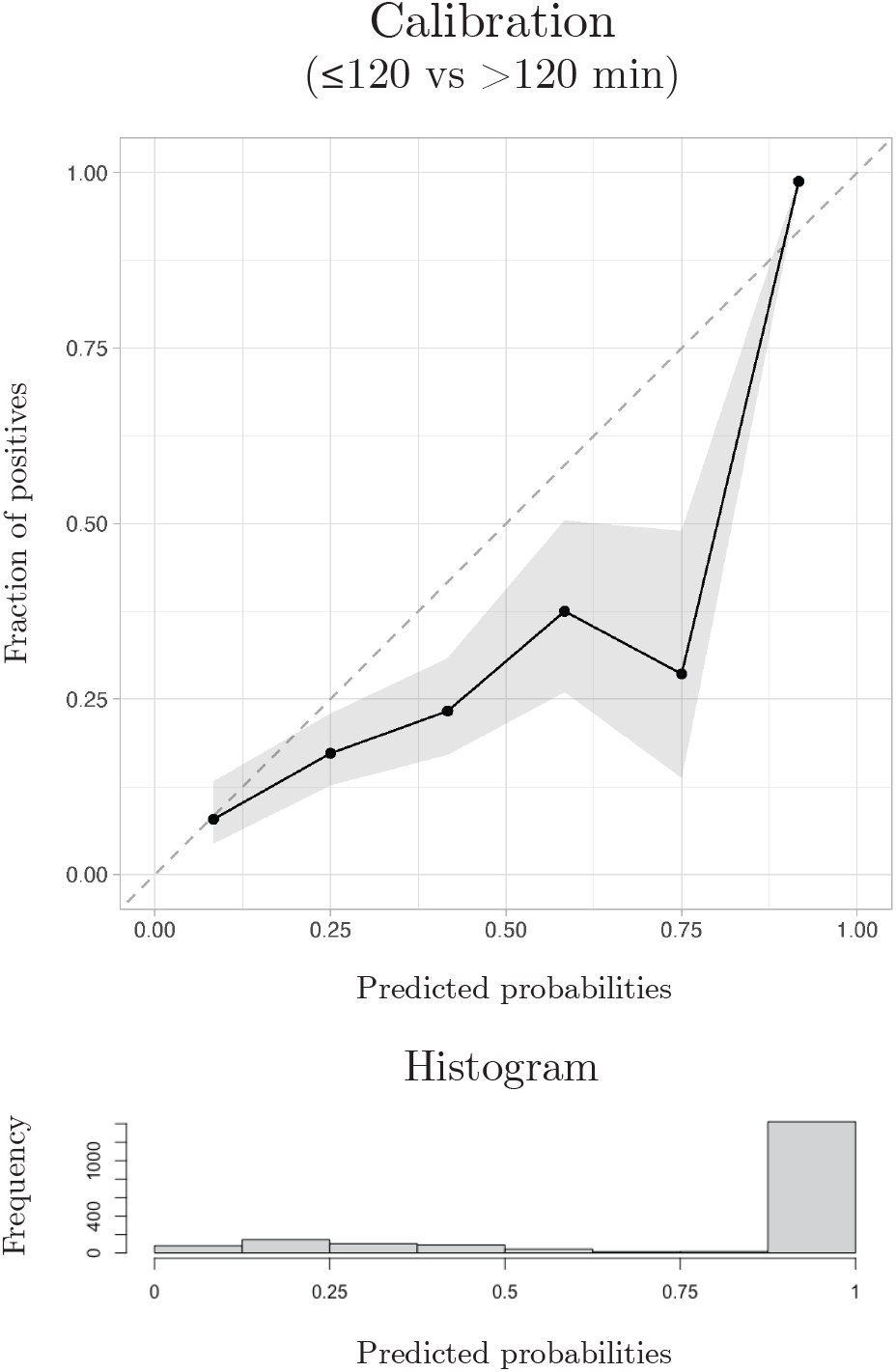
Calibration plot for the binary classification task TTD <120 min vs. TTD >120 min on the external validation cohort, computed with the R package val.prob.ci.2. The predicted probabilities come from the output of our model and plotted against the fraction of positives observed in the data. The histogram shows the prevalence of patients for different ranges of predicted probabilities.

